# Upper airway gene expression differentiates COVID-19 from other acute respiratory illnesses and reveals suppression of innate immune responses by SARS-CoV-2

**DOI:** 10.1101/2020.05.18.20105171

**Authors:** Eran Mick, Jack Kamm, Angela Oliveira Pisco, Kalani Ratnasiri, Jennifer M. Babik, Carolyn S. Calfee, Gloria Castañeda, Joseph L. DeRisi, Angela M. Detweiler, Samantha Hao, Kirsten N. Kangelaris, G. Renuka Kumar, Lucy M. Li, Sabrina A. Mann, Norma Neff, Priya A. Prasad, Paula Hayakawa Serpa, Sachin J. Shah, Natasha Spottiswoode, Michelle Tan, Stephanie A. Christenson, Amy Kistler, Charles Langelier

## Abstract

We studied the host transcriptional response to SARS-CoV-2 by performing metagenomic sequencing of upper airway samples in 238 patients with COVID-19, other viral or non-viral acute respiratory illnesses (ARIs). Compared to other viral ARIs, COVID-19 was characterized by a diminished innate immune response, with reduced expression of genes involved in toll-like receptor and interleukin signaling, chemokine binding, neutrophil degranulation and interactions with lymphoid cells. Patients with COVID-19 also exhibited significantly reduced proportions of neutrophils and macrophages, and increased proportions of goblet, dendritic and B-cells, compared to other viral ARIs. Using machine learning, we built 26-, 10- and 3-gene classifiers that differentiated COVID-19 from other acute respiratory illnesses with AUCs of 0.980, 0.950 and 0.871, respectively. Classifier performance was stable at low viral loads, suggesting utility in settings where direct detection of viral nucleic acid may be unsuccessful. Taken together, our results illuminate unique aspects of the host transcriptional response to SARS-CoV-2 in comparison to other respiratory viruses and demonstrate the feasibility of COVID-19 diagnostics based on patient gene expression.

## Introduction

The emergence of severe acute respiratory syndrome coronavirus 2 (SARS-CoV-2) in December 2019 has precipitated a global pandemic with over 4.5 million cases and 300,000 deaths^1^. Coronavirus disease 2019 (COVID-19), the clinical syndrome caused by SARS-CoV-2, varies from asymptomatic infection to critical illness, with dysregulated inflammatory response to infection a hallmark of severe cases^2^. Defining the host response to SARS-CoV-2, as compared to other respiratory viruses, is fundamental to identifying mechanisms of pathogenicity and potential therapeutic targets.

Metagenomic next generation RNA sequencing (mNGS) is a powerful tool for assessing host/pathogen dynamics^3,4^ and a promising modality for developing novel respiratory diagnostics that integrate host transcriptional signatures of infection^3,5^. While proven for diagnosis of other acute respiratory infections^3,5^, transcriptional profiling has not yet been evaluated as a diagnostic tool for COVID-19, despite its potential to mitigate the risk of false negatives associated with standard naso/oropharyngeal (NP/OP) swab-based PCR testing^6–8^.

## Results and Discussion

To interrogate the molecular pathogenesis of SARS-CoV-2 and evaluate the feasibility of a COVID-19 diagnostic based on host gene expression, we conducted a multicenter observational study of 238 patients with acute respiratory illnesses (ARIs) who were tested for SARS-CoV-2 by NP/OP swab PCR, and performed host/viral mNGS on the same specimens. The cohort (**Table S1**) included 94 patients who tested positive for SARS-CoV-2 by PCR, 41 who tested negative but had other pathogenic respiratory viruses detected by mNGS (**Methods, Figure S1A**), and 103 with no virus detected (non-viral ARIs).

We began by performing pairwise differential expression analyses between the three patient groups (**Methods, Supp. File 1**). Hierarchical clustering of the union of the 50 most significant genes in each of the comparisons revealed that the transcriptional response to SARS-CoV-2 was distinct from the response to other viruses (**Figure 1A**). We detected gene clusters that were up- (cluster I) or down-regulated (cluster II) by other viruses as compared to non-viral ARIs, but relatively unaffected by SARS-CoV-2. Importantly, we also identified a small number of genes that were upregulated by SARS-CoV-2 more than by other viruses (cluster III). And many genes upregulated in all viral ARIs (cluster IV) appeared to respond to SARS-CoV-2 proportionally to viral load, as measured by the relative abundance of sequencing reads mapped to the virus (**Methods, Figure S1B**).

**Figure 1.**
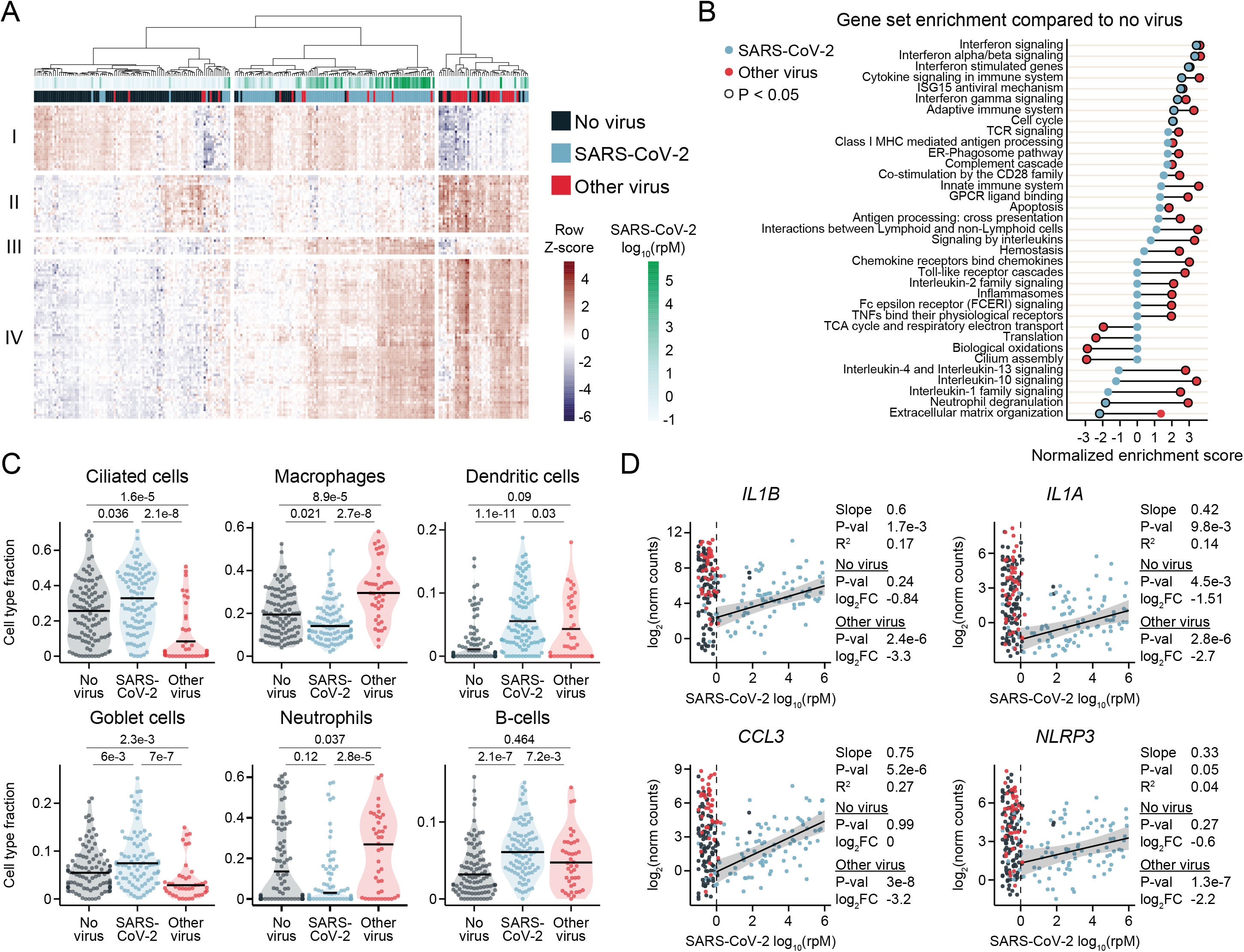
Host Transcriptional Signatures of SARS-CoV-2 Infection as Compared to Other Respiratory Viruses. **A**. Hierarchical clustering of 120 genes comprising the union of the top 50 DE genes by significance in each of the pairwise comparisons between patients with COVID-19 (SARS-CoV-2), other viral ARIs and non-viral ARIs. Group labels and viral load of SARS-CoV-2 are shown in the annotation bars. rpM, reads-per-million. **B**. Normalized enrichment scores of selected REACTOME pathways that achieved statistical significance (Benjamini-Hochberg adjusted p-value < 0.05) in at least one of the gene set enrichment analyses, using either DE genes between SARS-CoV-2 and non-viral ARIs or between other viruses and non-viral ARIs. If a pathway could not be tested in one of the comparisons since it had less than 10 members in the input gene set, the enrichment score was set to 0. **C**. *In silico* estimation of cell type fractions in the bulk RNA-seq using lung single cell signatures. Black lines denote the median. The y-axis in each panel was trimmed at the maximum value among the three patient groups of 1.5*IQR above the third quartile. All pairwise comparisons were performed with a two-sided Mann-Whitney-Wilcoxon test followed by Bonferroni’s correction. **D**. Scatter plots of normalized gene counts (log_2_ scale) as a function of SARS-CoV-2 viral load, log_10_ (rpM). Robust regression was performed on SARS-CoV-2 positive patients with log_10_(rpM) > 0 to highlight the relationship to viral load. Shown are inflammasome-related genes selected from among the genes most depressed in expression in SARS-CoV-2 compared to other viral ARIs. Statistical results for each gene refer to (from top to bottom): the regression analysis, the DE analysis between SARS-CoV-2 and non-viral ARIs, and the DE analysis between SARS-CoV-2 and other viral ARIs.

To investigate the pathways driving these distinctions, we performed gene set enrichment analyses^9^ (GSEA) on the genes differentially expressed (DE) between SARS-CoV-2 and non-viral ARIs, and separately, those DE between other viral ARIs and non-viral ARIs (**Methods, Supp. File 2**). We found that both SARS-CoV-2 and other viruses elicited an interferon response in the upper airway (**Figure 1B**). The most significant genes upregulated by SARS-CoV-2 were interferon inducible, including *IFI6, IFI44L, IFI27* and *OAS2* (**Figure S2A**), in agreement with previous reports^10,11^. *IFI27* was induced by SARS-CoV-2 significantly more than by other viruses, even at low viral load. Most other top DE genes, however, did not distinguish COVID-19 from other viral ARIs. *ACE2*, which encodes the cellular receptor for SARS-CoV-2, was also non-specifically induced, consistent with its recent identification as a general interferon stimulated gene^12^.

Notably, GSEA of DE genes in the direct comparison of SARS-CoV-2 and other viruses suggested elements of the interferon response to SARS-CoV-2 were attenuated (**Figure S2B, Supp. File 2**). Indeed, numerous interferon response genes, such as *IRF7* and *OASL*, were more potently induced by other viruses, and high SARS-CoV-2 abundance was required to achieve comparable induction (**Figure S2C**). These results may be related to observations of a blunted interferon response in cellular models of SARS-CoV-2 infection^13^, though the effects in patients appear more nuanced.

A striking contrast between SARS-CoV-2 and other viruses emerged in the activation of additional innate immune signaling pathways (**Figure 1B, S2B**). Other viral ARIs caused significant upregulation of gene expression associated with toll-like receptors, interleukin signaling, chemokine binding, neutrophil degranulation and interactions with lymphoid cells, yet the response of these pathways to SARS-CoV-2 was markedly attenuated (**Figure 1B, S2B**). While other viral ARIs appeared to depress expression of genes involved in cilia functions and antioxidant responses, this was not observed for SARS-CoV-2 (**Figure 1B, S2B**).

*In silico* estimation of cell type proportions revealed significant differences between the groups (**Figure 1C, S3**). Compared to patients with other viral and non-viral ARIs, those infected with SARS-CoV-2 exhibited significantly reduced fractions of monocytes/macrophages and neutrophils, and significantly increased proportions of goblet, dendritic and B-cells. Patients with other viral ARIs exhibited decreased ciliated cell and ionocyte fractions, and increased macrophage, neutrophil and T-cell fractions, compared to those with non-viral ARIs. These results closely aligned with the GSEA findings and suggested that the diminished innate immune responses in COVID-19 patients were coupled to differences in the cellular composition of the airway microenvironment.

The gene that was most decreased in expression in COVID-19 patients compared to those with other viral ARIs was *IL1B*, which encodes a pro-inflammatory cytokine produced by the inflammasome complex, particularly in macrophages^14^ (**Figure 1D, Supp. File 1**). Among the top 100 differentially decreased genes were those involved in inflammasome activation and activity *(NLRP3, CASP5, IL1A, IL1B, IL18RAP, IL1R2)* and in chemokine signaling for recruiting innate immune cells to the epithelium *(CCL2, CCL3, CCL4)*. Given that IL1-β and other pro-inflammatory cytokines are primary targets of monoclonal antibody therapeutics under investigation^15^, these results raise the question of whether further suppression early in the course of disease may be detrimental in the setting of an already suppressed inflammatory response to SARS-CoV-2.

Relatively few genes were specifically upregulated in COVID-19 patients compared to both other viral and non-viral ARIs. These included *TRO*, which encodes a membrane-bound cell adhesion molecule; *WDR74*, which plays a role in rRNA processing and associates with the RNA helicase MTR4^16^; *EIF4A2*, a translation initiation factor that has been shown to interact with other coronaviruses as well as HIV^17,18^; and *FAM83A*, which is involved in epidermal growth factor receptor (EGFR) signaling^19^.

We next asked whether host gene expression data could be used to construct a classifier capable of accurately differentiating COVID-19 from other ARIs (viral or non-viral). By employing a combination of lasso regularized regression and random forest (**Methods**), we first identified a 26-gene signature that performed with an area under the receiver operating characteristic curve (AUC) of 0.980 (range of 0.951-1.000), as estimated by 5-fold cross validation (**Figure 2A, Tables S2, S3**). Even though many patients undergoing testing for COVID-19 may not be infected with other respiratory viruses, we recognized the need for classifier specificity in this circumstance and examined how well the classifier performed at distinguishing SARS-CoV-2 from other respiratory viruses. We found that it achieved an AUC of 0.966 (range 0.895-1.000) when tested only on patients with other viral ARIs, indicating robust specificity for SARS-CoV-2 (**Tables S2, S3**). Using a cut-off of 40% predicted out-of-fold probability for COVID-19 to call a patient positive, this translated into a sensitivity of 97% and a specificity of 96% for patients with non-viral ARIs and 83% for patients with other viral ARIs (**Figure 2B**).

**Figure 2.**
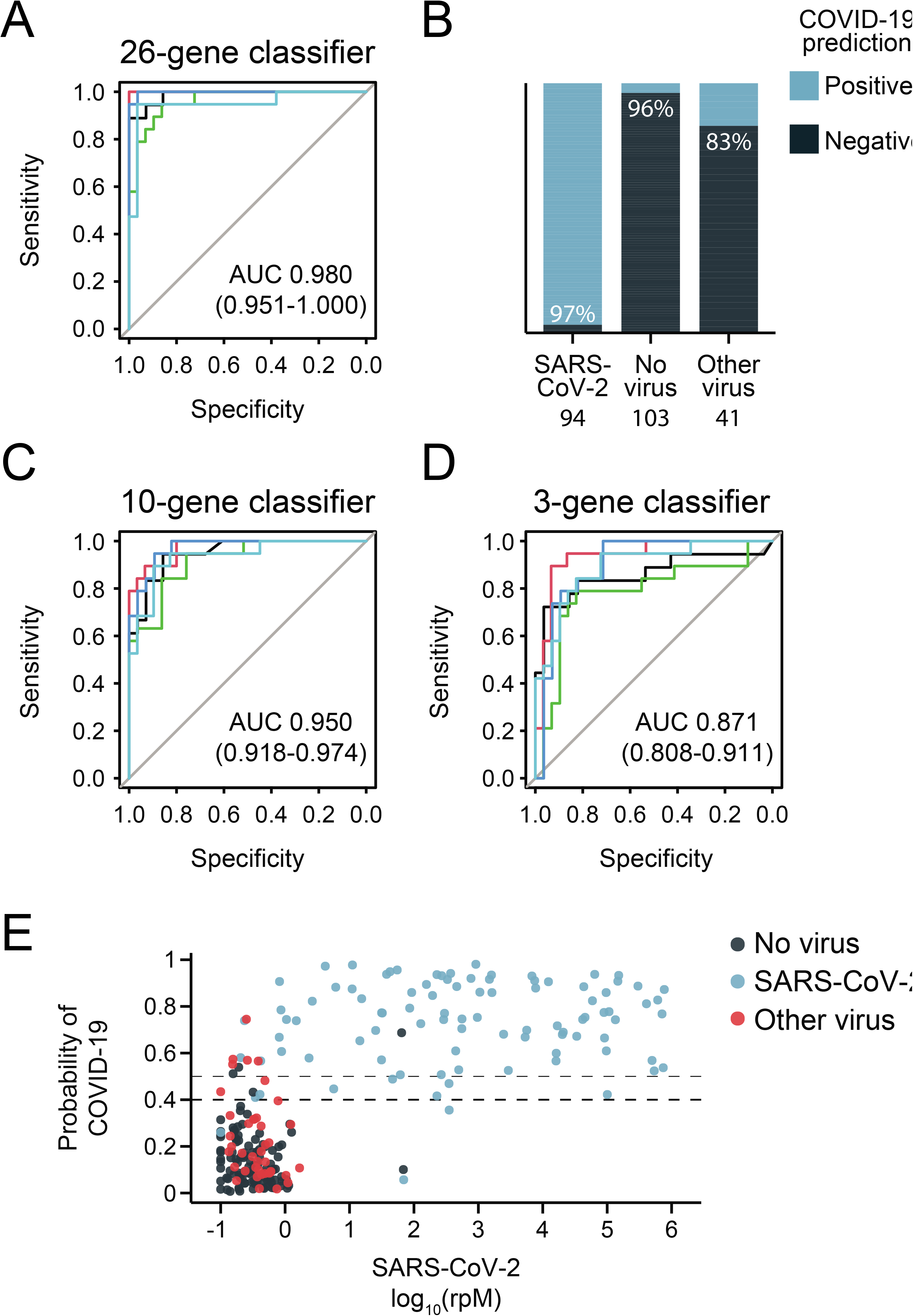
Performance of COVID-19 Diagnostic Classifiers Based on Patient Gene Expression. **A**. Receiver operating characteristic (ROC) curve for a 26-gene classifier that differentiates COVID-19 from other acute respiratory illnesses (viral and non-viral). **B**. Accuracy of the 26-gene classifier within each patient group, using a cut-off of 40% out-of-fold predicted probability for COVID-19. **C**. ROC curve for a 10-gene classifier. **D**. ROC curve for a 3-gene classifier. **E**. Out-of-fold predicted probability of COVID-19 derived from the 26-gene classifier plotted as a function of SARS-CoV-2 viral load, log_10_ (rpM). Dashed lines indicate 40% (our chosen cut-off) and 50%.

Given that a parsimonious gene set could enable practical incorporation into a clinical PCR assay, we implemented a more restrictive regression penalty and identified a 10-gene classifier that could differentiate SARS-CoV-2 from other respiratory illnesses with an AUC of 0.950 (range 0.918-0.974) (**Figure 2C, Tables S2, S3**). Classification performance specifically against other viral ARIs suffered slightly but still achieved an AUC of 0.905 (range 0.842-0.959). Existing SARS-CoV-2 PCR assays typically employ 3 gene targets and thus we tested the potential to further reduce host classifier gene size. We found that a sparse 3-gene *(IL1B, IFI6, IL1R2)* classifier still achieved an AUC of 0.871 (range 0.808-0.911) (**Figure 2D, Tables S2, S3**).

A host-based diagnostic might have particular utility if it could increase the sensitivity of standard NP/OP swab PCR testing, which may return falsely negative in a significant proportion of patients^6–8^. Presumably, false negatives are in large part due to insufficient viral abundance in the collected specimen. While our cohort did not include PCR-negative samples from patients with clinically confirmed COVID-19, we evaluated whether classifier performance was affected by viral load. The predicted probability of SARS-CoV-2 infection had little apparent relationship to the abundance of SARS-CoV-2, suggesting host gene expression has the potential to provide an orthogonal indication of COVID-19 status even when viral abundance is low (**Figure 2E**).

In summary, we studied 238 patients with acute respiratory illnesses to define the human upper respiratory tract gene expression signature of COVID-19. Our study is limited by sample size, incomplete demographic data and the need for an independent validation cohort. Notwithstanding, our results illuminate unique aspects of the host transcriptional response to SARS-CoV-2 in comparison to other respiratory viruses and provide insight regarding molecular pathogenesis. We also leveraged these data to develop an accurate, clinically practical, COVID-19 diagnostic classifier that may help overcome the limitations of direct detection of viral nucleic acid. Future prospective studies in a larger cohort will be needed to validate these findings, determine the prognostic value of host signatures, and assess the performance of integrated host/viral diagnostic assays.

## Data Availability

Gene counts, sample metadata, and code to generate viral calls by mNGS, perform DE, regression and cell type analyses, and construct the gene expression classifiers are available at:

https://github.com/czbiohub/covid19-transcriptomics-pathogenesis-diagnostics-results

## Materials and Methods

### Study design, clinical cohort and ethics statement

We conducted an observational cohort study of patients with acute respiratory illnesses suspected to be COVID-19 at the University of California, San Francisco (UCSF) and Zuckerberg San Francisco General Hospital between 03/10/2020 and 04/07/2020. Through UCSF IRB protocol 17-24056, a waiver of consent was granted to evaluate unused clinical specimens in the UCSF Clinical Microbiology Laboratory and assess demographics and basic clinical features from the Epic-based electronic health record.

### SARS-CoV-2 detection by clinical PCR

Testing for COVID-19 was carried out in the UCSF Clinical Microbiology Laboratory using polymerase chain reaction (PCR) of NP swab or pooled NP + OP swab specimens using primers targeting either two regions of the SARS-CoV-2 N gene (n=156, 66%), or the E and RNA-dependent RNA polymerase genes (n=82, 34%), plus human RNAse P as a positive control. In all our analyses, we defined patients with COVID-19 as those with a positive SARS-CoV-2 result by PCR.

### Metagenomic sequencing

To evaluate host gene expression and detect the presence of other viruses, metagenomic next generation sequencing (mNGS) of RNA was performed on the same specimens subjected to SARS-CoV-2 PCR testing. Following DNase treatment, human cytosolic and mitochondrial ribosomal RNA was depleted using FastSelect (Qiagen). To control for background contamination, we included negative controls (water and HeLa cell RNA) as well as positive controls (spike-in RNA standards from the External RNA Controls Consortium (ERCC))^1^. RNA was then fragmented and subjected to a modified metagenomic spiked sequencing primer enrichment (MSSPE) library preparation method^2^. Briefly, a 1:1 mixture of the NEBNext Ultra II RNAseq Library Prep (New England Biolabs) random primers and a pool of SARS-CoV-2 primers at 100 μM was used at the first strand synthesis step of the standard RNAseq library preparation protocol to enrich for reads spanning the length of the SARS-CoV-2 genome. RNA-seq libraries underwent 146 nucleotide paired-end Illumina sequencing on an Illumina Novaseq 6000 instrument.

### Quantification of SARS-CoV-2 abundance by mNGS

All samples were processed through a SARS-CoV-2 reference-based assembly pipeline that involved removing non-SARS-CoV-2 reads with Kraken2^3^, and aligning to the SARS-CoV-2 reference genome MN908947.3 using minimap2^4^. We calculated SARS-CoV-2 reads-per-million (rpM) using the number of reads that aligned with mapq >= 20. For plotting purposes, 0.1 was added to the rpM values to avoid taking the log of 0.

### Detection of other respiratory pathogenic viruses by mNGS

All samples were processed through the IDSeq pipeline^5,6^, which performs reference based alignment at both the nucleotide and amino acid level against sequences in the National Center for Biotechnology Information (NCBI) nucleotide (NT) and non-redundant (NR) databases, followed by assembly of the reads matching each taxon detected. We further processed the results for viruses with established pathogenicity in the respiratory tract^3^. We evaluated whether one of these viruses was present in a patient sample if it met the following three initial criteria: i) at least 10 counts mapped to NT sequences, ii) at least 1 count mapped to NR sequences, iii) average assembly nucleotide alignment length of at least 70bp.

Negative control (water and HeLa cell RNA) samples enabled estimation of the number of background reads expected for each virus, which were normalized by input mass as determined by the ratio of sample reads to spike-in positive control ERCC RNA standards^7^. Viruses were then additionally tested for whether the number of sequencing reads aligned to them in the NT database was significantly greater compared to negative controls. This was done by modeling the number of background reads as a negative binomial distribution, with mean and dispersion fitted on the negative controls. For each batch (sequencing run) and taxon (virus), we estimated the mean parameter of the negative binomial by averaging the read counts across all negative controls after normalizing by ERCCs, slightly regularizing this estimate by including the global average (across all batches) as an additional sample. We estimated a single dispersion parameter across all taxa and batches, using the functions glm.nb() and theta.md() from the R package MASS^8^. We considered a patient to have a respiratory pathogenic virus detected by mNGS if the virus achieved an adjusted p-value < 0.05 after Holm-Bonferroni correction for all tests performed in the same sample.

### Host differential expression (DE) analysis

Following demultiplexing, sequencing reads were pseudo-aligned with kallisto^9^ (v. 0.46.1; including bias correction) to an index consisting of all transcripts associated with human protein coding genes (ENSEMBL v. 99), cytosolic and mitochondrial ribosomal RNA sequences, and the sequences of ERCC RNA standards. Samples retained in the dataset had a total of at least 400,000 estimated counts associated with transcripts of protein coding genes, and the average across all samples was 5.79 million. Gene-level counts were generated from the transcript-level abundance estimates using the R package tximport^10^, with the lengthScaledTPM method.

Genes were retained for differential expression analysis if they had at least 10 counts in at least 20% of samples (n=15,900). The analysis was performed with the R package limma^11^ using quantile normalization and the design: ~0 + viral status + gender + age + sequencing batch, where viral status was either “SARS-CoV-2”, “other virus” or “no virus”. We note that the gender of patients for whom we lacked this information was inferred based on chromosome Y gene expression, and the age of patients for whom we lacked this information was taken as the mean age of samples with age reported in the respective viral status group.

To generate the gene expression heatmap, hierarchical clustering was performed on the union of the top 50 genes (by p-value) in each of the pairwise comparisons among the three groups (n=120 genes). Gene counts were subjected to the variance stabilizing transformation, as implemented in the R package DESeq2^12^, centered and scaled prior to clustering. For both rows and columns, Euclidean distance was the distance measure and Ward’s criterion (ward.D2) was the agglomeration method.

### Gene set enrichment analysis

Gene set enrichment analyses^13^ were performed using the fgseaMultilevel function in the R package fgsea^14^ on REACTOME^15^ pathways with a minimum size of 10 genes and a maximum size of 1,000. The genes included in each pairwise comparison were those with Benjamini-Hochberg adjusted p-value < 0.1 and |log_2_(FC)| > log(1.5) in the respective DE analysis, preranked by fold change.

The gene sets shown in Figure 1B were manually selected to reduce redundancy and highlight diverse biological functions from among those with a Benjamini-Hochberg adjusted p-value < 0.05 in at least one of the comparisons i) SARS-CoV-2 vs. no virus, and ii) other virus vs. no virus. And the gene sets shown in Figure S2B were similarly selected from among those with an adjusted p-value < 0.05 in the direct comparison of SARS-CoV-2 vs. other virus. Full results of all analyses are provided as supplementary.

### Regression of gene counts against viral abundance

We performed robust regression of the limma-generated quantile normalized gene counts against log_10_ (rpM) of SARS-CoV-2 for all genes with a Benjamini-Hochberg adjusted p-value < 0.001 in either the DE analysis of SARS-CoV-2 vs. no virus, or SARS-CoV-2 vs. other virus (n=2,920). The samples included were those in the SARS-CoV-2 patient group with rpM >= 1. Robust regression was used to better account for outlier data points.

The analysis was performed using the R package robustbase^16^, which implements MM-type estimators for linear regression^17,18^, using the KS2014 setting and the model: quantile normalized counts (log_2_ scale) ~ gender + age + sequencing batch + log_10_ (rpM). Model predictions for the log_10_ (rpM) co-variate were used for display in the individual gene plots. Reported p-values for significance of the difference of the regression coefficient from 0 were Benjamini-Hochberg adjusted, and reported R^2^ values represent the adjusted robust coefficient of determination^19^.

### *In silico* analysis of cell type fractions

Cell-type fractions were estimated from bulk host transcriptome data using the CIBERSORT X algorithm^20^. We used the human lung cell atlas dataset^21^ to derive the single cell signatures. The cell types estimated with this reference cover all expected cell types in the airway samples. The estimated fractions were compared between the three patient groups using a two-sided Mann-Whitney-Wilcoxon test with Bonferroni correction.

### Classifier construction

We built sparse classifiers for COVID-19 status using a combined lasso and random forest approach. For feature selection, we used the logistic lasso (as implemented in the R package glmnet^22^), and then trained random forests on the selected features (using the R package randomForest^23^). We used 5-fold cross-validation to evaluate model error. For each train-test split, we used a nested cross-validation within the training set to select the lasso tuning parameter. For the random forest, we used 10,000 trees, and left all tuning parameters at their defaults. For the initial input features (before selection), we used gene counts with a variance-stabilizing transform derived from the training set only (using the R package DESeq2^12^). Classifiers were built using a gold standard of COVID-19 diagnosis based on SARS-CoV-2 PCR positivity.

### Data availability

Gene counts, sample metadata, and code to generate viral calls by mNGS, perform DE, regression and cell type analyses, and construct the gene expression classifiers are available at: https://github.com/czbiohub/covid19-transcriptomics-pathogenesis-diagnostics-results

## Supplementary Tables

**Table S1.**
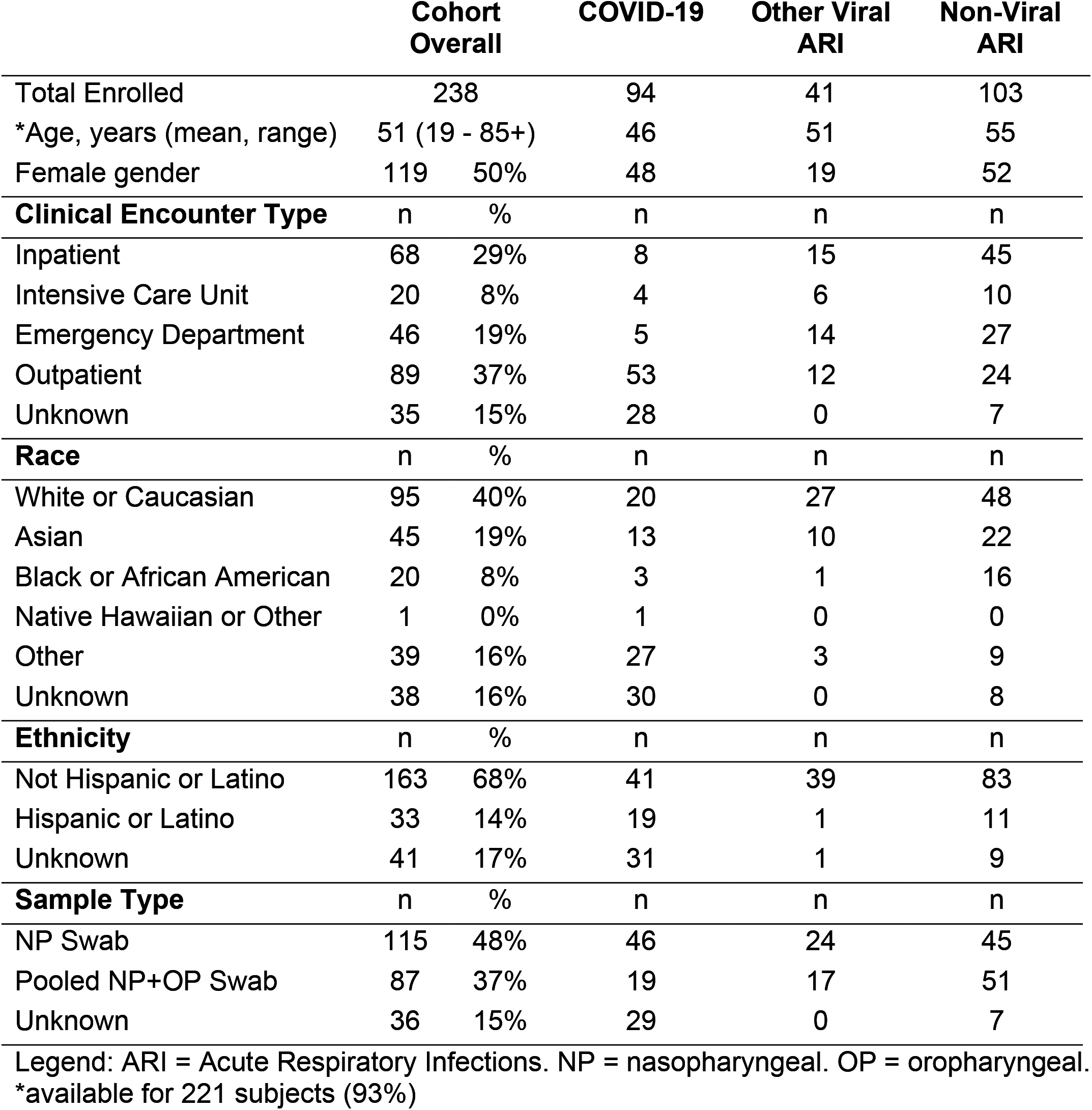
Cohort Clinical and Demographic Characteristics.

|  | **Cohort Overall** |  | **COVID-19** | **Other Viral ARI** | **Non-Viral ARI** |
| --- | --- | --- | --- | --- | --- |
| Total Enrolled | 238 |  | 94 | 41 | 103 |
| *Age, years (mean, range) | 51 (19 - 85+) |  | 46 | 51 | 55 |
| Female gender | 119 | 50% | 48 | 19 | 52 |
| **Clinical Encounter Type** | n | % | n | n | n |
| Inpatient | 68 | 29% | 8 | 15 | 45 |
| Intensive Care Unit | 20 | 8% | 4 | 6 | 10 |
| Emergency Department | 46 | 19% | 5 | 14 | 27 |
| Outpatient | 89 | 37% | 53 | 12 | 24 |
| Unknown | 35 | 15% | 28 | 0 | 7 |
| **Race** | n | % | n | n | n |
| White or Caucasian | 95 | 40% | 20 | 27 | 48 |
| Asian | 45 | 19% | 13 | 10 | 22 |
| Black or African American | 20 | 8% | 3 | 1 | 16 |
| Native Hawaiian or Other | 1 | 0% | 1 | 0 | 0 |
| Other | 39 | 16% | 27 | 3 | 9 |
| Unknown | 38 | 16% | 30 | 0 | 8 |
| **Ethnicity** | n | % | n | n | n |
| Not Hispanic or Latino | 163 | 68% | 41 | 39 | 83 |
| Hispanic or Latino | 33 | 14% | 19 | 1 | 11 |
| Unknown | 41 | 17% | 31 | 1 | 9 |
| **Sample Type** | n | % | n | n | n |
| NP Swab | 115 | 48% | 46 | 24 | 45 |
| Pooled NP+OP Swab | 87 | 37% | 19 | 17 | 51 |
| Unknown | 36 | 15% | 29 | 0 | 7 |

Legend: ARI = Acute Respiratory Infections. NP = nasopharyngeal. OP = oropharyngeal.
\*available for 221 subjects (93%)

**Table S2.** A. Performance of classifier models.

| **Model** | **COVID-19 vs.**<br>**All Other ARI** | **COVID-19 vs.**<br>**Non-viral ARI** | **COVID-19 vs.**<br>**Other Viral ARI** |
| --- | --- | --- | --- |
| 26-gene | 0.980 (0.951-1.000) | 0.985 (0.947-1.000) | 0.966 (0.895-1.000) |
| +age/gender | 0.970 (0.936-0.991) | 0.974 (0.932-1.000) | 0.959 (0.914-0.994) |
| 10-gene | 0.950 (0.918-0.974) | 0.968 (0.945-0.997) | 0.905 (0.842-0.959) |
| 3-gene | 0.871 (0.808-0.911) | 0.930 (0.893-0.969) | 0.722 (0.539-0.842) |

| **Model** | **Threshold** | **Accuracy** | **Sensitivity** | **Specificity** |
| --- | --- | --- | --- | --- |
| 26-gene | 0.5 | 0.924 | 0.894 | 0.944 |
| 10-gene | 0.5 | 0.882 | 0.851 | 0.903 |
| 3-gene | 0.5 | 0.836 | 0.766 | 0.882 |
| 26-gene | 0.4 | 0.941 | 0.968 | 0.924 |
| 10-gene | 0.4 | 0.878 | 0.883 | 0.875 |
| 3-gene | 0.4 | 0.832 | 0.809 | 0.847 |

**Table S3.**
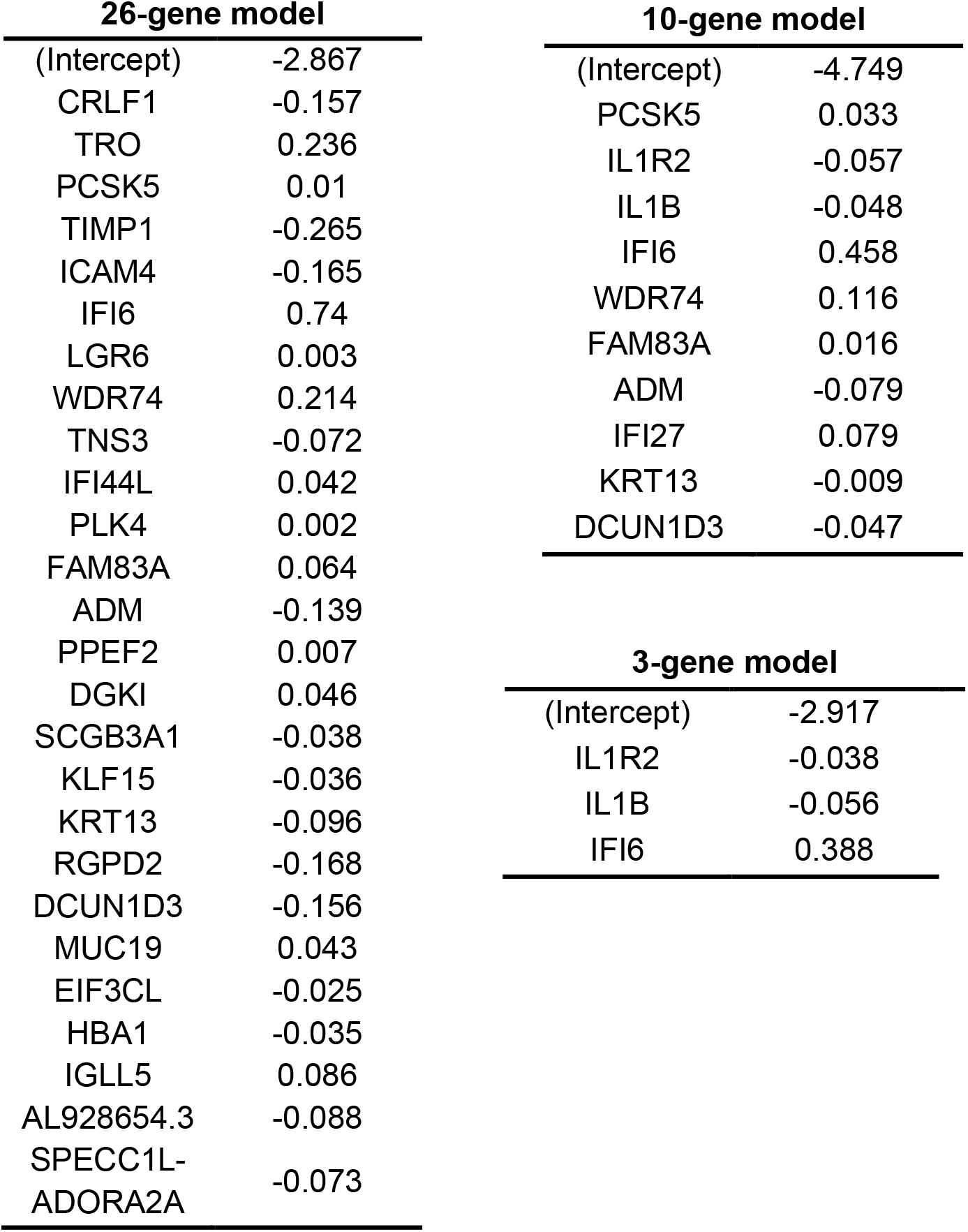
Lasso-selected features and coefficients of classifier models.

## Supplementary Files

**Supplementary File 1**. Differential expression analyses.

**Supplementary File 2**. Gene set enrichment analyses.

**Supplementary File 3**. Cell type fractions.

**Supp. Figure 1.**
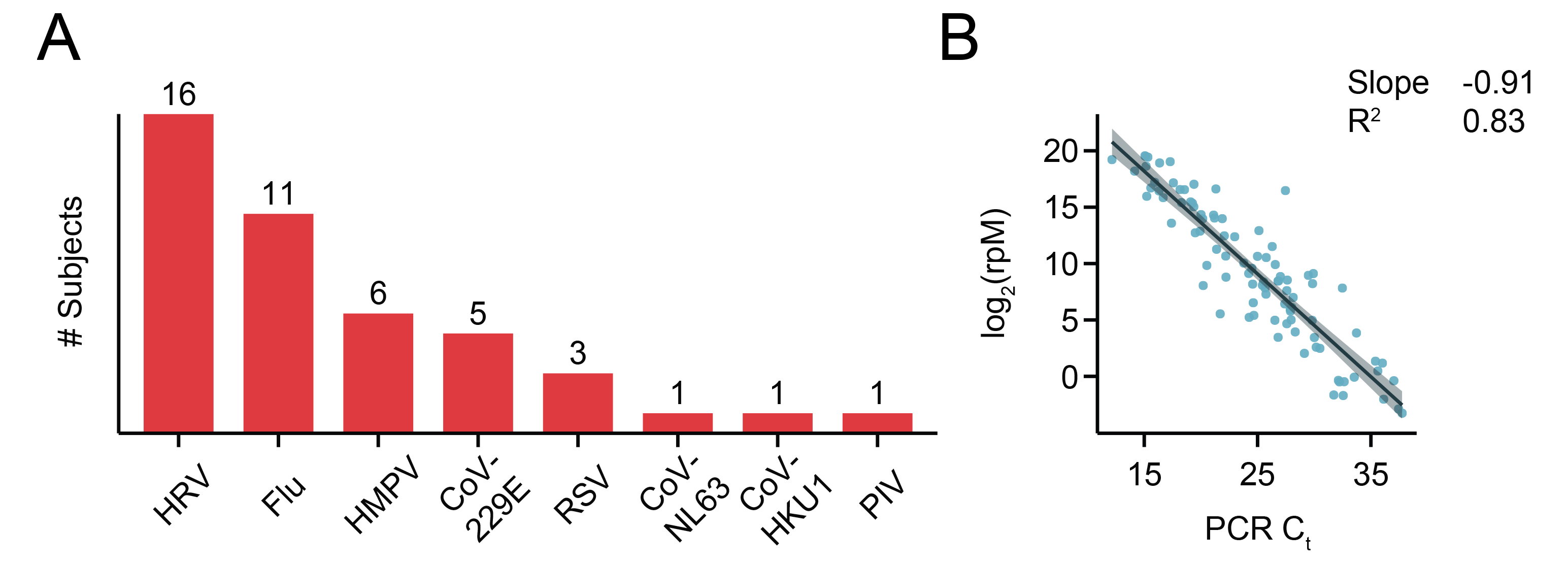
**A**. Breakdown of subjects with other pathogenic respiratory viruses identified by mNGS. Three patients had viral/viral co-infections: SARS-CoV-2/HRV (n=1) and RSV/HRV (n=2). CoV=Coronavirus, HRV=Human Rhinovirus, Flu=Influenza Virus, HMPV=Human Metapneumovirus, RSV=Respiratory Syncytial Virus, PIV=Parainfluenza Virus. **B**. Correlation of SARS-CoV-2 PCR Crossing Threshold (Ct) and mNGS reads-per-million (rpM). Ct represents an average across the SARS-CoV-2 genomic loci assessed.

**Supp. Figure 2.**
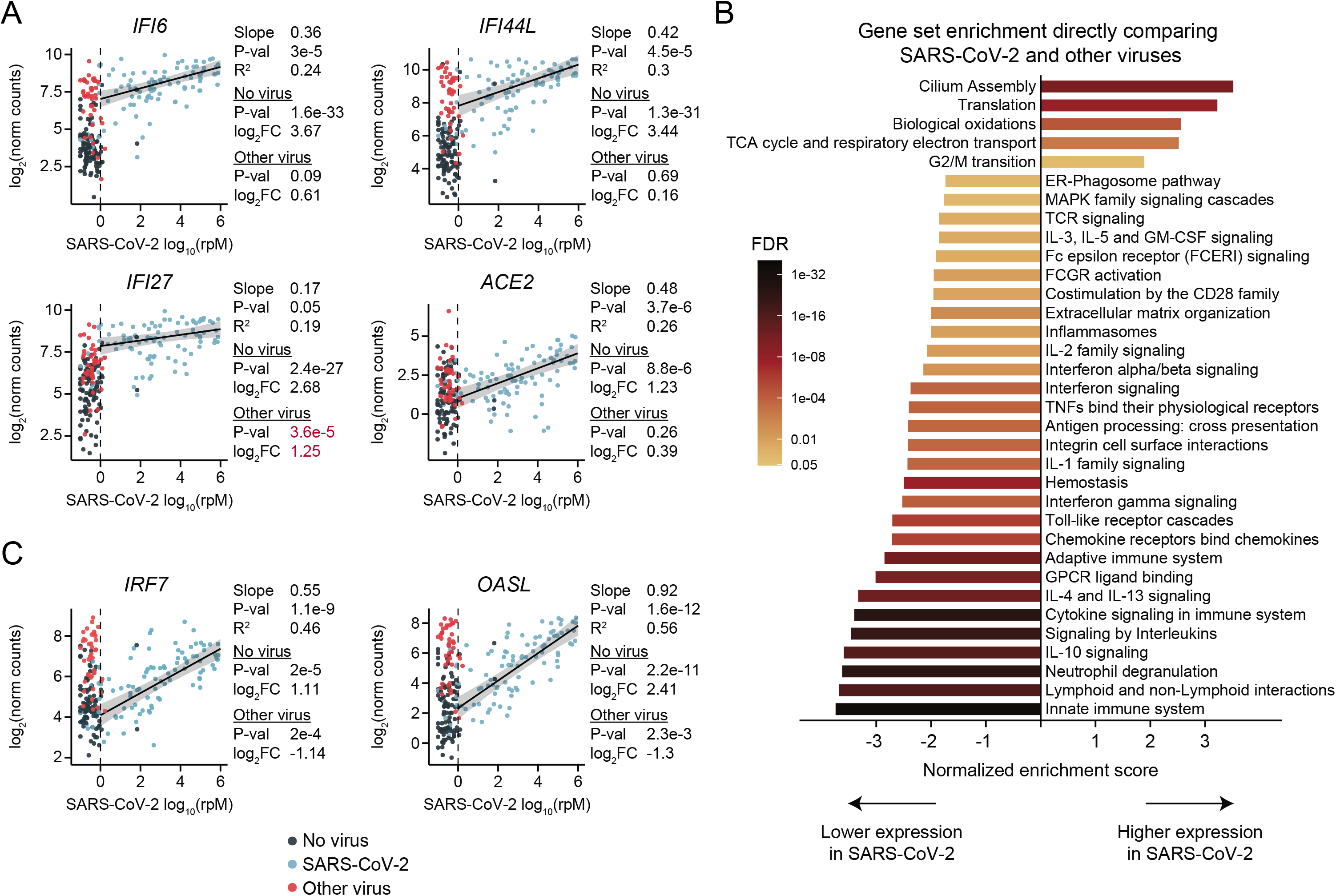
**A**. Gene expression scatter plots for the most significant interferon response genes induced by SARS-CoV-2, and the SARS-CoV-2 receptor gene ACE2. **B**. Gene set enrichment analysis for the direct comparison between COVID-19 and other viral ARIs. **C**. Gene expression scatter plots for selected interferon response genes in the leading edge of the interferon signaling gene set, showing lagging expression in SARS-CoV-2 compared to other viral ARIs.

**Supp. Figure 3.**
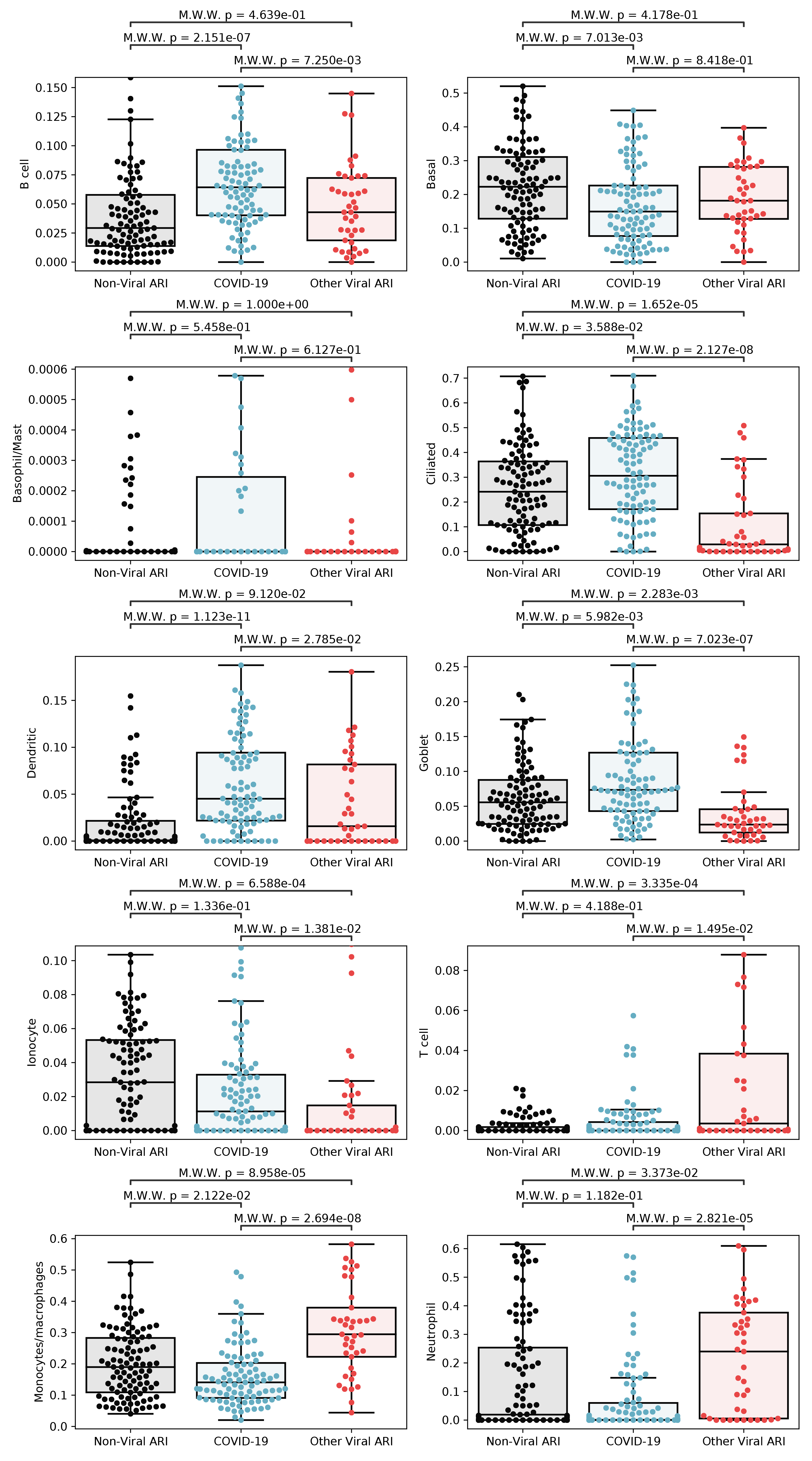
*In silico* estimation of cell type fractions in the bulk RNA-seq using lung single cell signatures. Black lines denote the median. The y-axis in each panel was trimmed at the maximum value among the three patient groups of 1.5*IQR above the third quartile. All pairwise comparisons were performed with a two-sided Mann-Whitney-Wilcoxon test followed by Bonferroni’s correction.

